# Pasteurisation temperatures effectively inactivate influenza A viruses in milk

**DOI:** 10.1101/2024.05.30.24308212

**Authors:** Jenna Schafers, Caroline J. Warren, Jiayun Yang, Junsen Zhang, Sarah J. Cole, Jayne Cooper, Karolina Drewek, B Reddy Kolli, Natalie McGinn, Mehnaz Qureshi, Scott M. Reid, Thomas P. Peacock, Ian Brown, Joe James, Ashley C. Banyard, Munir Iqbal, Paul Digard, Ed Hutchinson

## Abstract

In late 2023 an H5N1 lineage of high pathogenicity avian influenza virus (HPAIV) began circulating in American dairy cattle^1^. Concerningly, high titres of virus were detected in cows’ milk, raising the concern that milk could be a route of human infection. Cows’ milk is typically pasteurised to render it safe for human consumption, but the effectiveness of pasteurisation on influenza viruses in milk was uncertain. To assess this, we evaluated heat inactivation in milk for a panel of different influenza viruses. This included human and avian influenza A viruses (IAVs), an influenza D virus that naturally infects cattle, and recombinant IAVs carrying contemporary avian or bovine H5N1 glycoproteins. At pasteurisation temperatures, viral infectivity was rapidly lost and became undetectable before the times recommended for pasteurisation. We then showed that an H5N1 HPAIV in milk was effectively inactivated by a comparable treatment, even though its genetic material remained detectable. We conclude that industry standard pasteurisation conditions should effectively inactivate H5N1 HPAIV in cows’ milk, but that unpasteurised milk could carry infectious influenza viruses.

## Introduction

Since 2020 an H5N1 Clade 2.3.4.4b lineage of high pathogenicity influenza virus (HPAIV) has spread rapidly around the world, causing the worst outbreak of avian influenza on record^2,3^. H5N1 IAVs can cause severe disease in humans^4^ so the pandemic potential of this outbreak is of great concern^5^. While HPAIVs are able to cross between host species, viral adaptation to sustained transmission within mammalian populations is uncommon. The current H5N1 virus has caused repeated spillover infections in mammals, but most of these were in wild animals and not in close proximity to humans^6–8^. This changed in early 2024, when it was realised that H5N1 HPAIVs were spreading among dairy cattle in the USA^1^. This was alarming because of the extensive human-animal interface of the dairy industry, including the widespread consumption of dairy products. It was also surprising, for two reasons. Firstly, cattle had previously been considered resistant to IAV infection, with only sporadic cases reported^9,10^. Secondly, although IAV typically spreads by respiratory or faecal-oral transmission, H5N1 HPAIV was shed at startlingly high titres into milk^11^. Shedding into milk may have led to further spillover events on dairy farms, with H5N1 identified in dead farm cats, wild raccoons and foxes, cattle-associated perching birds, and nearby poultry flocks. Furthermore, HPAIV in cows has also resulted in at least two cases of conjunctivitis in dairy farm workers^12,13^ This new route of transmission has also resulted in H5N1 HPAIV being shed into milk sold for human consumption, with viral genetic material detected in as much as 20% of supermarket milk in some affected areas^14^. Determining if humans could be exposed to infectious H5N1 HPAIV through consuming cows’ milk is a matter of urgent importance.

Because cows’ milk can carry a variety of pathogens, it is typically pasteurised before human consumption. Pasteurisation is a well-established method of heat inactivation, which was first formalised by Pasteur for wine in 1864^15^ and which correlated with drastic falls in infant mortality and other diseases when widely applied to milk over the first half of the twentieth century^16,17^. It is assumed that pasteurisation of milk would also be effective against bovine H5N1 HPAIV, but this was based on general assumptions about the structure of the virus and very limited studies of heat treatment of other influenza viruses suspended in other substances ^18–21^. Encouragingly, initial reports indicated that infectious virus could not be recovered from pasteurised milk containing viral genetic material^22^, but without a general understanding of how influenza viruses in milk respond to pasteurisation it was hard to predict the robustness of commercial pasteurisation against this new strain of virus. Here, we address this by determining the general response of influenza viruses to pasteurisation temperatures in milk. As the consumption of unhomogenised and unpasteurised (‘raw’) milk is also popular in some affected areas, we also assessed whether influenza viruses remain infectious in milk if heating is not applied.

## Results

To assess the effects of pasteurising temperatures on influenza viruses, we first tested the responses of a variety of strains at biosafety containment level 2 (Table 1)^23^. We also used reverse genetics to generate a panel of 6:2 reassortant influenza viruses carrying the ‘internal’ genes of the laboratory strain A/Puerto Rico/8/1934 (PR8) and the surface proteins (HA and NA) of various representatives of H5N1 clade 2.3.4.4b, all ‘de-engineered’ to replace the polybasic cleavage site that renders them highly pathogenic with a monobasic cleavage sites (Table 2)^24^. For all viruses, we assessed the effect of pasteurising temperatures applied for specific time intervals by mixing virus 1:10 with milk and heating small volumes in thin-walled PCR tubes. The milk was then rapidly cooled, diluted in tissue culture medium and infectivity was assessed by plaque assay.

**Table 1:** Influenza viruses used in the study.

| Strain name | Short Name | Details |
| --- | --- | --- |
| A/Puerto Rico/8/1934 (H1N1) | PR8<br>(PR8:PB) | Laboratory strain<br>(PR8 refers to data collected at the Roslin Institute, and PR8:PB |
|  |  | to data collected at the Pirbright Institute) |
| BrightFlu | BF-PR8 | A PR8 derivative encoding a fluorescent marker (data collected at the MRC-University of Glasgow Centre for Virus Research)<sup>23</sup> |
| A/wild-duck/Italy/17VIR6926-1/2017 H5N2 (H5N2) | H5N2 | low pathogenicity avian influenza virus |
| A/Duck/Singapore/97 (H5N3) | H5N3 | low pathogenicity avian influenza virus<sup>44</sup> |
| D/bovine/France/5920/2014 | IDV | a separate genus of influenza virus that that naturally infects cattle |
| A/chicken/Scotland/054477/2021 | H5N1 | high pathogenicity avian influenza virus |

**Table 2:** Reassortant influenza viruses uses in the study.

| Source of HA and NA | Short Name | Details of HA and NA |
| --- | --- | --- |
| A/chicken/Scotland/054477/2021 | PR8:AIV09 | AIV09 (AB genotype) |
| A/chicken/England/085598/2022 | PR8:AIV48 | AIV48 (BB genotype) |
| A/dairy cow/Texas/24-008749-001-original/2024 | PR8:Cattle | cattle isolate |
| A/goat/Minnesota/24-007234-003-original/2024 | PR8:Goat | goat isolate |

Reassortant viruses contain an NA and a 'de-engineered' HA from the strain indicated, with the remaining genes from the laboratory strain PR8.

We chose temperatures representing the two common methods of pasteurising milk: low-temperature long time (LTLT; the vat method), which requires heating to 62.5⍰ for at least 30 minutes^25^; and high-temperature short time (HTST), which requires heating to 72⍰ for at least 15 seconds^26^. Our aim was not to test specific models of pasteurisation equipment, but rather to determine how quickly inactivation of influenza viruses occurred at the temperatures required for a well-conducted pasteurisation.

We first tested PR8 in both raw milk and commercially available whole milk (‘processed milk’), observing similar, rapid inactivation on heating in both cases (**Figure 1a**). We therefore tested our remaining panel of viruses in processed milk, choosing heating times that allowed us to assess the rate of inactivation (**Figure 1b,c**). At both 63⍰ and 72⍰ the infectivity of all viruses was rapidly lost, dropping by orders of magnitude in seconds and falling below the limit of detection well in advance of the minimum times required for milk pasteurisation.

**Figure 1.**
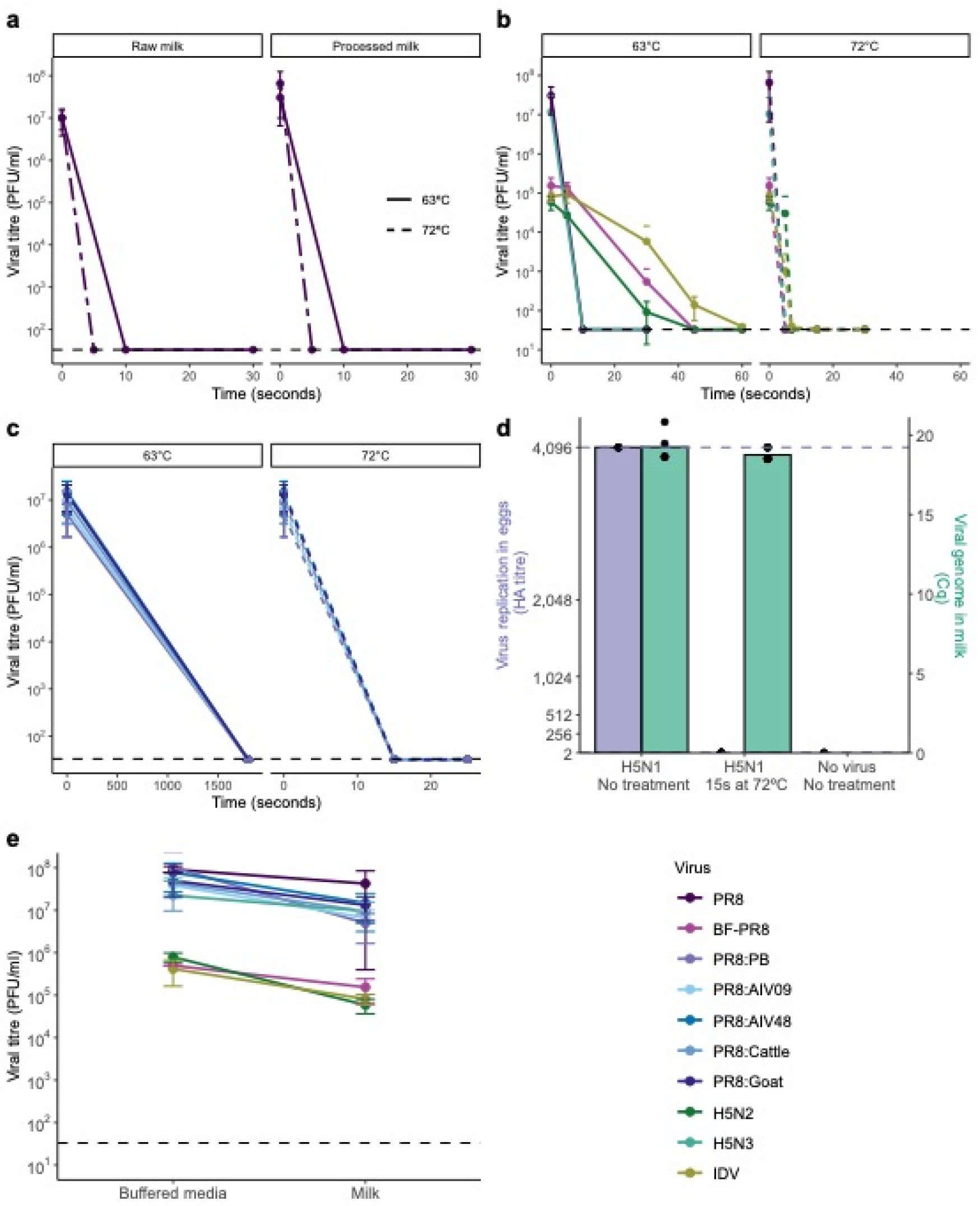
Pasteurisation effectively inactivates influenza viruses in milk. **(a)** PR8 was mixed with raw milk or ‘shop-bought’ pasteurised whole milk (‘processed milk’), heated for the indicated time and then cooled. Infectivity was measured by plaque assay. Data are the mean ± SD of two to three independent repeats, each done in duplicate. Limit of detection (LoD) = 33 PFU/ml. **(b)** Viruses were mixed with processed milk and treated as in (a). Mean ± SD of two to three independent repeats are shown. For BF-PR8, H5N2 and IDV LoD = 20 PFU/ml, for PR8 and H5N3 LoD= 33 PFU/ml. **(c)** Reassortant PR8 viruses with the HA and NA of the indicated strains were treated as in (a). Mean ± SD of three repeats are shown. Error bars without a lower limit represent a minimum below 0. **(d)** H5N1 HPAIV was mixed with raw milk, either unheated or pre-heated to 71.7°C, then cooled after 15s and used to inoculate three replicate eggs. Viral replication in eggs was assessed by haemagglutination assay (upper and lower LoD are 2^12^ and 2^1^ HAU, respectively). Viral genome in milk was detected using the H5 HPAIV rRT-PCR assay. The means of three repeats are shown along with, for each repeat, the individual Cq values of the milk and the mean HA titres of three replicate eggs. **(e)** Comparison of the plaque titres of influenza viruses when mixed with tissue culture medium/phosphate buffered saline, or with milk. Mean ± SD are shown, N varies from 2 – 8 depending on the condition. Error bars without a lower limit represent a minimum below 0. Details of viruses are given in Tables 1 and 2.

The fact that all influenza viruses responded similarly (including IDV, which has been reported to be unusually thermally stable^27^), strongly suggested that H5N1 IAV would also be susceptible to pasteurisation. To test this directly, we used the wild-type H5N1 strain A/chicken/Scotland/054477/2021 (AIV09/AB genotype) and mimicked the conditions of HTST pasteurisation at SAPO containment level 4. In this experiment raw milk, either unheated or pre-heated to 71.7°C, was mixed with one part in 100 of virus (final titre of 3×10^7^ EID_50_). After 15s the mixture was cooled on ice; viral genome was detected by RT-PCR and infectivity was assessed by inoculation of milk into embryonated fowls’ eggs (EFEs), followed by incubation and a haemagglutination assay of the allantoic fluid (**Figure 1d**). Heat treatment did not affect the detection of viral genomes in milk but, although infectious virus was isolated from room temperature milk, no infectious virus could be reproducibly isolated in EFEs following exposure to HTST pasteurisation conditions, either during direct inoculation (**Figure 1d**) or when inoculated material was passaged to a second EFE (data not shown). We conclude that heating to pasteurisation temperatures effectively inactivates influenza A and D viruses, including H5N1 HPAIVs, within the times required for pasteurisation.

Finally, we addressed the question of whether raw milk can carry infectious virus. We found that mixing influenza viruses with unheated milk caused some reduction in infectivity, consistent with previous studies^10^ (**Figure 1e**). However, it is important to note that this was never more than a moderate effect. For all viruses tested, including H5N1 HPAIV and PR8 with H5N1 surface proteins, unpasteurised milk was clearly an effective carrier of infectious influenza viruses (**Figure 1b-d**).

## Discussion

In this study we responded to reports that H5N1 HPAIV had been detected in milk from infected dairy cattle in the USA, by asking if pasteurisation of cows’ milk could inactivate influenza viruses. Given the urgency of this question we made two decisions in designing our study which should be considered when interpreting our results. Firstly, we made a general assessment of the times needed to inactivate influenza viruses by pasteurisation under well-controlled laboratory conditions. This allowed us to establish general principles which could be used for quality control assessments of specific industrial pasteurisation apparatuses. Secondly, as well as testing the effects of pasteurisation on a recent H5N1 HPAIV, we considered a panel of influenza viruses, including an influenza D virus with a potentially higher thermal tolerance^27^. This allowed us to establish general conditions for the inactivation of any influenza virus in milk by pasteurisation.

Overall, we found that pasteurisation temperatures of both 63⍰ (LTLT) and 72⍰ (HTST) rapidly and effectively inactivated influenza viruses in milk (**Figure 1b-d**). In the case of H5N1 HPAIV, treatment at 72⍰ eliminated infectivity without affecting the detection of viral genetic material, consistent with initial reports from the USA that detected viral genetic material but no infectious virus in pasteurised milk^11,14^ (**Figure 1d**). Our data also suggest the likely effects of thermal inactivation of influenza viruses in other situations (consistent with reports that beef spiked with H5N1 HPAIV and cooked to at least 62.5°C showed complete viral inactivation^28^), although direct testing of these other methods would still be advisable.

As this study was being prepared for publication, another study was published that took milk directly from cows infected with H5N1 HPAIV and tested viral inactivation at 63⍰ and 72⍰^29^. It is useful to compare the results of these studies. Importantly, the main findings are consistent – at either temperature, when infectious virus was quantified in cell culture systems the inactivation kinetics were (as far as can be judged from the timepoints tested) very similar, showing a rapid loss of orders of magnitude of infectivity and with the limits of detection reached before the minimum times required for pasteurisation. This was observed despite minor variations between the equipment and methods used across multiple laboratories in the two studies (compare for example the different experiments with the PR8 strain noted in Table 1). However, in one important difference from our work the other study reported that, for milk that had been heated at 72⍰ for up to 30s, some residual infectivity could be detected by inoculation into EFEs. This is close to the lower time limits of HTST pasteurisation, though as with our study their work was performed in a laboratory setting rather than with industrial pasteurisation equipment.

The reason for the difference between our studies requires further investigation, but the data available suggest two explanations. The authors of the other study noted that foot-and-mouth disease virus has previously been found to be more heat stable when shed into milk by an infected animal than when spiked in experimentally^30,31^, although it is not yet clear if influenza viruses gain a similar thermal protection when shed naturally into milk. A second explanation for the difference is suggested by the inactivation kinetics described by our data. Thermal inactivation of influenza viruses is not instantaneous, and slight differences in inactivation conditions can shift the time at which the virus becomes completely undetectable (**Figure 1b,c**). Overall, this highlights an important area for further work: although the infectious dose for oral infection by HPAIV H5N1 in milk is not yet known, more work would be needed to precisely define the shortest heat treatment that could completely eliminate infectivity. For now, the inactivation time courses we present here can be considered when determining if a pasteurisation process takes milk well past the point where infectious influenza viruses should be recoverable.

Finally, although our data are encouraging regarding the safety of pasteurised milk that has been contaminated with H5N1 HPAIV, they also highlight that without pasteurisation milk can carry infectious influenza virus, a finding that has also been confirmed by others^29^. We therefore caution against the consumption of raw milk that could be contaminated with bovine IAV because of the risk of consuming infectious virus, in addition to its established risk for infection with other viral and bacterial pathogens^16,25^.

## Materials and Methods

### Cells and Viruses

For work at biosafety containment level 2, PR8 and BrightFlu were generated by reverse genetics, as previously described^32^. These viruses, as well as A/Duck/Singapore/97 (H5N3) (a gift of Prof Wendy Barclay, Imperial College) and A/wild-duck/Italy/17VIR6926-1/2017 (H5N2) (a gift of Dr Isabella Monne, Istituto Zooprofilattico Sperimentale delle Venezie) were propagated on Madin Darby Canine Kidney carcinoma (MDCK) cells, while D/bovine/France/5920/2014 (IDV, a gift of Dr Mariette Ducatez, Université de Toulouse) was propagated on Swine Testis (ST) cells. To generate reassortant viruses, HA and NA sequences were synthesised by GenScript and cloned into the pHW2000 vector. The polybasic cleavage site of H5 HA was replaced by a monobasic site to allow the work to be conducted at biosafety containment level 2. Viruses were rescued using the pHW2000 eight-plasmid bidirectional expression system^33^ with the internal segments from PR8. Reassortant viruses were propagated in 9 to 10-day old embryonated fowls’ eggs to generate working stocks. The GISAID accession numbers of the strains used for the reassortant viruses are: EPI_ISL_9012696, EPI_ISL_13782459, EPI_ISL_19014384 and EPI_ISL_19015123.

Work at SAPO containment level 4 used A/chicken/Scotland/054477/2021, an H5N1-2021 clade 2.3.4.4 HPAIV derived from an UK outbreak event and representative of the UK/European epizootic season in 2021. The virus was propagated in 9 to 10-day-old specified-pathogen free embryonated eggs.

### Pasteurisation Assays

For work at biosafety containment level 2, virus stocks were diluted 1:10 (v/v) in test solutions. These were either ‘buffered solutions’ (phosphate buffered saline (PBS) or DMEM), or milk. Milk used was either ‘processed’ (homogenised and pasteurised whole milk, purchased from supermarkets in the United Kingdom, which at the time of writing has no confirmed cases of bovine IAV) or ‘raw’ (obtained directly from cows in a herd managed by the University of Edinburgh, and used without prior processing). Milk was either used on the day of acquisition, or kept refrigerated or frozen at –20°C to prevent spoilage prior to experimentation. To test heat inactivation, 100μl of diluted virus was aliquoted into 200μl thin-walled PCR strip tubes (ThermoFisher), with sealed lids to prevent evaporation. These were placed in a thermocycler, exposed to either 72 or 63°C for a set time period, then rapidly cooled and placed on ice. The thermocycler lid was typically heated to the same temperature as the block, or higher, to limit condensation. Thermocycler models used were an Applied Biosystems Veriti™ 96-Well Fast Thermal Cycler (Roslin Institute), and BIO RAD T100™ (MRC-University of Glasgow Centre for Virus Research, Pirbright Institute).

For work at SAPO containment level 4, 3×10^9^ EID_50_ units of virus were mixed 1:100 (v/v) into unpasteurised whole milk (1 ml final volume with a final titre of 3×10^7^ EID^50^), either at room temperature or pre-heated in a hot block to 71.7 °C, and then placed on ice after 15s.

### Virus titration

For work at biosafety containment level 2, virus infectivity was determined by plaque assay in MDCK cells after dilution in tissue culture medium (this was necessary as undiluted milk had a pronounced cytopathic effect). Plaques were visualised either by direct staining of the monolayer or, in the case of IDV, labelled by immunocytochemistry with a custom sheep polyclonal antibody against IDV NP (available from www.influenza.bio; third bleed used at 1/500), an Alexa Fluor™ 568 donkey anti-sheep secondary (Thermo) and a DAPI counterstain, and visualised with a Celigo imaging cytometer (Nexcelom).

For work at SAPO containment level 4, Cq values were determined using an H5 HP rRT-PCR assay^34^, and infectivity of the allantoic fluid of inoculated specified-pathogen free embryonated fowls’ eggs was determined by haemagglutination assay.

### Analysis

Data processing, analysis and visualisation was performed using the R statistical computing software in R Studio (version 2023.06.0+421)^35–37^. Figures were produced using packages ggplot2 and ggpubr^38,39^. Other packages included RMisc^40^, scales^41^, janitor^42^ and ggh4x^43^. The data and materials necessary to reproduce the findings and figures reported are available at the Open Science Framework (https://osf.io/m4fa5).

## Data Availability

All data produced in the present study are available upon reasonable request to the authors.

## Acknowledgments

We acknowledge support for this research consortium from the Medical Research Council (MRC), Biotechnology and Biological Sciences Research Council (BBSRC) and Department for Environment, Food and Rural Affairs (Defra, UK) as ‘FluMAP’ [grant number BB/X006204/1, BB/X006166/1], ‘FluTrailMap’ [grant number BB/Y007271/1, BB/Y007298/1] and FluTrailMap-One Health [MR/Y03368X/1]. We also acknowledge funding from the MRC to E.H. [MC_PC_21023 for the Influenza Virus Toolkit and MC_UU_00034/1 to the MRC-University of Glasgow Centre for Virus Research] and from the BBSRC to P.D. [Institute Strategic Programme grant BBS/E/RL/230002D and Evolution & Ecology of Infectious Disease grant BB/V011286/1]. J.S. is supported by an Edinburgh Clinical Academic Track fellowship from the Wellcome Trust, and the Centre for Open Science via Flu lab. APHA staff were funded by the UK Department for the Environment, Food and Rural Affairs (Defra) and the devolved Scottish and Welsh governments under grants SE2213, SV3400 and SV3006. The Pirbright Institute staff are funded by the BBSRC via Institute Strategic Programme Grants (ISPGs) [BBS/E/PI/230002A, BBS/E/PI/230002B]. We thank Professor Alastair Macrae (Royal (Dick) School of Veterinary Science) for providing raw cows’ milk, Dr M Khalid Zakaria (MRC-University of Glasgow Centre for Virus Research) for assistance with preparing virus stocks, and the staff of the MRC Protein Phosphorylation and Ubiquitylation Unit, University of Dundee for their assistance in antibody generation. We gratefully acknowledge all data contributors, i.e., the authors and their originating laboratories responsible for obtaining the specimens, and their submitting laboratories for generating the genetic sequence and metadata and sharing via the GISAID Initiative, for the genomic data on which this research is based. All submitters of the data may be contacted directly via the GISAID website (https://www.gisaid.org).

## Author Contributions

J.S. conceptualisation, methodology, investigation, writing – original draft, writing – review and editing, visualisation, data curation; J.Z. conceptualisation, methodology, investigation, writing – review and editing; J.Y. conceptualisation, methodology, investigation, writing – review and editing; C.J.W. investigation; S.J.C. methodology; M.Q. conceptualisation, investigation; B.R.K investigation; J.C. investigation; K.D. investigation; N.McG. investigation; S.M.R investigation; J.J. supervision; A.C.B. writing – review and editing, supervision; T.P.P. resources, writing – review and editing, supervision, funding acquisition; I.B. writing – review and editing, supervision, funding acquisition; M.I. writing – review and editing, supervision, funding acquisition; P.D. writing – review and editing, supervision, funding acquisition; E.H. conceptualisation, writing – original draft, writing – review and editing, supervision, funding acquisition.

